# Triglyceride-glucose index and the risk of heart failure: evidence from two large cohorts and a Mendelian randomization analysis

**DOI:** 10.1101/2022.06.28.22277033

**Authors:** Xintao Li, Jeffrey Shi Kai Chan, Bo Guan, Shi Peng, Xiaoyu Wu, Jiandong Zhou, Jeremy Man Ho Hui, Yan Hiu Athena Lee, Danish Iltaf Satti, Shek Long Tsang, Shouling Wu, Songwen Chen, Gary Tse, Shaowen Liu

**Author notes:** Corresponding author: Professor Shaowen Liu, MD, PhD, Department of Cardiology, Shanghai General Hospital, School of Medicine, Shanghai Jiao Tong University, 200080 Shanghai, China, Co-corresponding author: Professor Gary Tse, MD, PhD, Kent and Medway Medical School, Canterbury, Kent, CT2 7NT, United Kingdom., Dr. Songwen Chen, MD, PhD, Department of Cardiology, Shanghai General Hospital, School of Medicine, Shanghai Jiao Tong University, 200080 Shanghai, China. These authors have contributed equally to this work and share first authorship (Xintao Li, Jeffrey Shi Kai Chan).

## Abstract

**Background:** The relationship between triglyceride-glucose (TyG) index, an emerging marker of insulin resistance, and the risk of incident heart failure (HF) was unclear. This study thus aimed to investigate this relationship.

**Methods:** Subjects without prevalent cardiovascular diseases from the prospective Kailuan cohort (recruited during 2006-2007) and a retrospective cohort of family medicine patients from Hong Kong (recruited during 2000-2003) were followed up until December 31^st^, 2019 for the outcome of incident HF. Separate adjusted hazard ratios (aHRs) summarizing the relationship between TyG index and HF risk in the two cohorts were combined using a random-effect meta-analysis. Additionally, a two-sample Mendelian randomization (MR) of published genome-wide association study data was performed to assess the causality of observed associations.

**Results:** In total, 95,996 and 19,345 subjects from the Kailuan and Hong Kong cohorts were analyzed, respectively, with 2,726 cases (2.8%) of incident HF in the former and 1,709 (7.0%) in the latter. Subjects in the highest quartile of TyG index had the highest risk of incident HF in both cohorts (Kailuan: aHR 1.23 (95% confidence interval: 1.09-1.39), *P_Trend_* <0.001; Hong Kong: aHR 1.21 (1.04-1.40), *P_Trend_* =0.007; both compared with the lowest quartile). Meta-analysis showed similar results (highest versus lowest quartile: HR 1.22(1.11-1.34), *P*<0.0001). Findings from MR analysis, which included 47,309 cases and 930,014 controls, supported a causal relationship between higher TyG index and increased risk of HF (odds ratio 1.27(1.15-1.40), *P*<0.001).

**Conclusion:** A higher TyG index is an independent and causal risk factor for incident HF in the general population.

**Clinical Perspective:** *What is new?:*

- In 115,341 subjects from two large cohorts in China, an elevated triglyceride-glucose (TyG) index was independently associated with an increased risk of incident heart failure (HF).
- Two-sample Mendelian randomization analysis based on published genome-wide association studies found significant association between genetically determined TyG index and the risk of HF.
- Together, these findings suggest that a higher TyG index is an independent and causal risk factor of incident HF in the general population.

*What are the clinical implications?:*

- The TyG index may facilitate recognition of individuals at elevated risk of incident HF and allow early preventive interventions.
- The demonstrated causal effect of TyG index on incident HF warrants further research to fully understand the underlying mechanisms.

## Introduction

Heart failure (HF) is associated with significant morbidity and mortality, with contemporary five-year survival rates of less than 50%^1^. The prevalence of HF has been estimated to be 1-2% in developed countries and is projected to double by 2060^2, 3^. Given the enormous public health and socioeconomic burden caused by HF, it is critically important to identify individuals at high risk of HF and to implement preventive interventions as early as possible^4^.

Recently, the role of metabolic disorders in the development of HF has been increasingly investigated^5^. Insulin resistance, a hallmark of type II diabetes mellitus and metabolic syndrome, has been observed to be associated with adverse cardiac remodeling and dysfunction^6^. Molecular studies have provided ample evidence for the etiological role of insulin resistance in the development of HF^7, 8^. However, the gold standard method for measuring insulin sensitivity, the hyperinsulinaemic-euglycaemic clamp test, is time-consuming and invasive^9^, which has impeded its widespread use in clinical practice.

The triglyceride-glucose (TyG) index, a simple, dimensionless marker derived from fasting blood triglyceride and glucose levels as measured in routine biochemical tests, has been proposed and validated as a surrogate marker of insulin resistance^10^. Previous studies have found a positive association between TyG index and the risk of various metabolic and atherosclerotic cardiovascular diseases^11, 12^. However, few studies have been conducted to investigate the association between TyG index and the risk of incident HF, and whether the association is causal remains undetermined.

Mendelian randomization (MR) makes use of genetic variants as instrumental variables (IVs) to generate causal estimates of the long-term effects of risk factors on outcomes^13^. MR analysis can overcome the limitations of residual confounding and reverse causation in conventional observational studies^13, 14^. With the development of genome-wide association studies (GWAS), MR is highly suited and has been used for investigating the causal association between TyG index and HF^15, 16^.

As such, the present study aimed to assess the association between the TyG index and the risk of incident HF, as well as using a two-sample MR study to determine whether such associations were causal in nature.

## Methods

### Study design and population

Study subjects were identified from two Chinese studies, the Kailuan cohort in northern China and a territory-wide cohort in Hong Kong. The protocol for this study was in accordance with the guidelines of the Helsinki Declaration and this study was approved by the Ethics Committee at the Kailuan General Hospital and the Institutional Review Board of the University of Hong Kong / Hospital Authority Hong Kong West Cluster.

The Kailuan Study is a prospective cohort that based on a community in the Tangshan City. Details of the study has been published elsewhere^17^. In brief, a total of 101,510 subjects (aged 18 – 98 years; 81,110 males) were enrolled in the Kailuan Study at baseline (2006-2007), and received an interview of standardized questionnaires and clinical examinations at 11 hospitals responsible for health care of the community. The subjects were then followed up with repeat questionnaires, clinical and laboratory examinations every two years. All subjects gave informed consent to their enrolment in this study. Subjects with prevalent cancer and cardiovascular diseases, including HF, atrial fibrillation (AF), myocardial infarction (MI), and ischemic stroke were excluded, as well as those with missing baseline levels of triglyceride (TG) or fasting blood glucose (FBG).

Data for the Hong Kong cohort were extracted retrospectively from the Clinical Data Analysis and Reporting System (CDARS), an administrative electronic medical records database that records the basic demographics, diagnoses, selected procedures, medication prescriptions, and selected laboratory measurements of all patients that attended public healthcare institutions in Hong Kong which serve an estimated 90% of the population^18^. Diagnoses in CDARS were recorded using International Classification of Diseases, Ninth revision (ICD-9) codes regardless of the time of data entry, as ICD-10 has not been implemented in CDARS to date. The ICD-9 codes used for identifying comorbid conditions and the outcome (HF) were summarized in **Table S1**. CDARS has been extensively used in prior studies and has been shown to have good diagnostic coding accuracy^19, 20^. As only retrospective, deidentified data were used, the requirement for individual patient consent has been waived. For this study, adult patients (18 years old or above) attending a family medicine clinic in Hong Kong during the years 2000-2003 with at least one set of paired FBG and fasting TG levels at baseline were included. Patients with a history of ischemic heart disease, stroke, HF, AF, or cancer were excluded, as well as those who were pregnant at the time of inclusion, and those with missing baseline low-density lipoprotein cholesterol (LDL-C), high-density lipoprotein cholesterol (HDL-C), and total cholesterol levels.

### Data collection and definitions

The data collected and definitions used in this study were detailed in **Supplementary Methods**^17, 21, 22^. The TyG index was calculated as ln [fasting TG (mg/dl)×FBG (mg/dl) / 2]^23^.

### Outcomes and follow-up

In the Kailuan cohort, all subjects were followed from the baseline examination until the date of onset of HF, date of death, or end of follow-up (December 31^st^, 2019), whichever came first. HF was diagnosed by experienced cardiologists in accordance with the guidelines of the European Society of Cardiology^24^. Incident HF cases were derived from the Municipal Social Insurance Institutions, hospital discharge register, and death certificates.

In the Hong Kong cohort, all patients were followed up from inclusion until the first recorded diagnosis of HF, death, or the end of follow-up (December 31^st^, 2019), whichever came first. HF was identified using ICD-9 codes as summarized in **Table S1**.

### Two-sample MR analysis

Mendelian randomization is built upon three main assumptions^25^. First, single-nucleotide polymorphisms (SNPs) selected as instrumental variables should be robustly associated with the risk factor. Second, no association should exist between genetic variants and confounders. Third, genetic variants should affect outcome only through the risk factor.

TyG index-associated variants that reached genome-wide significance (*P* < 5 × 10^-8^) were retrieved from a previous GWAS^16^. In brief, the identified GWAS included 273,368 subjects from the United Kingdom Biobank, who were aged 40-69 and free from diabetes mellitus and lipid metabolism disorders^16^. These SNPs were further pruned by linkage disequilibrium with R^2^ < 0.01 and those that were significantly associated with TG or glucose were also excluded. In total, 192 IVs were selected for TyG index initially. Summary statistics data for the associations of TyG index-associated SNPs with HF were extracted from the published GWAS performed by the Heart Failure Molecular Epidemiology for Therapeutic Targets (HERMES) Consortium on 47,309 cases and 930,014 controls of European ancestry^26^. HF cases from 26 cohorts of the HERMES Consortium were identified based on the clinical diagnosis of HF of any etiology with no specific criteria for left ventricular ejection fraction. Details of subject selection were published elsewhere^26^.

### Statistical analysis

Continuous variables were presented as mean ± standard deviation (SD) or median with interquartile range (IQR) depending on their distribution. Categorical variables were presented as frequencies and percentages.

Kaplan-Meier curves were used to visualize the cumulative incidence of HF across quartiles of the TyG index. The association between baseline TyG index and the risk of incident HF was analyzed using the Cox proportional hazards model, with hazard ratios (HR) with 95% confidence intervals (CI) as the summary statistics. The Cox regression was performed with a staged approach, as detailed in **Supplementary Methods.** The association between the risks of HF and the observed spectrum of TyG index was also modelled and visualized using fractional polynomial curves with full multivariable adjustments. Furthermore, competing risk regression using the Fine and Gray sub-distribution model was performed to address the potentially confounding issue of competing risk, with death from any cause as the competing event. Sub-hazard ratios (SHR) with 95% CI were used as the summary statistics. Sensitivity analyses were conducted by excluding subjects with less than two-year follow-up time, and, separately, those with medications at baseline.

*A priori* subgroup analyses were performed for age (<65 vs ≥65), gender (male vs female), diabetes (yes vs no), hypertension (yes vs no), dyslipidemia (yes vs no) for both cohorts, and, for the Kailuan cohort, for obesity (yes vs no), and hs-CRP level (<1 mg/dl vs ≥1 mg/dl).

To combine the results from the two cohorts, we extracted hazard ratios from the fully adjusted model and performed a meta-analysis using the inverse variance method with random effects to estimate the association between TyG index, both as categorical and continuous variables, and the risk of incident HF.

In the MR analysis, the summary exposure and outcome data were first harmonized, and SNPs significantly associated with incident HF were excluded. Causal effects of TyG index on HF were estimated by the inverse-variance weighted (IVW) method. Weighted median, MR-Egger, and pleiotropy residual sum and outlier (MR-PRESSO) methods were used for supplementary analyses. Directional pleiotropy was assessed by MR-Egger intercepts and heterogeneity among genetic variants was evaluated by Cochran’s *Q* test.

To test the validity of causal effects estimates, multivariable MR (MVMR) using the IVW method was conducted to further investigate the direct causal effect of TyG index on HF after adjusting for confounders including BMI^27^, SBP^28^, DBP^28^, LDL-c^29^, and HDL-c^29^. An additional sensitivity analysis was performed by excluding any SNP significantly associated with those confounders.

All statistical analyses for the Kailuan and Hong Kong cohorts were conducted using SAS version 9.4 (SAS Institute, Inc., Cary, NC), Stata 16.1 software (StataCorp, College Station, TX), and/or RevMan (Version 5.1; Cochrane Collaboration, Oxford, UK). The MR analyses were performed by the TwoSampleMR, MR-PRESSO and MVMR packages with R version 4.0.2. All p values were two-sided, with *p* < 0.05 considered statistically significant.

## Results

Of the 101,510 subjects who took part in the Kailuan study, 95,996 subjects were analyzed after applying the exclusion criteria (**Figure S1**). For the Hong Kong cohort, 24,338 patients were identified for inclusion, and 19,345 patients were analyzed after applying the exclusion criteria (**Figure S2**). **Table 1** and **Table 2** shows the baseline characteristics of subjects according to the baseline TyG index quartiles of two cohorts.

**Table 1.** The baseline characteristics of subjects in the Kailuan Cohort.

| Characteristics | Total | TyG index |  |  |  |
| --- | --- | --- | --- | --- | --- |
|  |  | Q1 | Q2 | Q3 | Q4 |
|  |  | 3.60-8.18 | 8.18-8.57 | 8.57-9.05 | 9.05-12.51 |
| Subjects (n) | 95,996 | 23,997 | 24,000 | 24,001 | 23,998 |
| TyG index | 8.65±0.69 | 7.85±0.27 | 8.38±0.11 | 8.79±0.14 | 9.58±0.46 |
| Age, years | 51.4±12.5 | 50.2±13.7 | 51.4±12.7 | 52.1±12.2 | 52.0±11.4 |
| Male, n (%) | 76,364(79.6) | 17,655(73.6) | 18,994(79.1) | 19,501(81.3) | 20,214(84.2) |
| Height (cm) | 167.4±7.0 | 166.7±7.0 | 167.5±7.0 | 167.6±7.0 | 167.9±6.9 |
| BMI (kg/m<sup>2</sup>) | 25.0±3.5 | 23.4±3.2 | 24.6±3.3 | 25.6±3.3 | 26.4±3.4 |
| Completed high school, n (%) | 18,746(19.5) | 5673(23.6) | 4483(18.7) | 4448(18.5) | 4142(17.3) |
| Income≥800¥, n (%) | 13,133(13.7) | 3563(14.9) | 3093(12.9) | 3234(13.5) | 3243(13.5) |
| Daily Smoker, n (%) | 28,675(29.9) | 6902(28.8) | 6741(28.1) | 7173(29.9) | 7859(32.8) |
| Daily alcohol user, n (%) | 16,725(17.4) | 3891(16.2) | 3834(16.0) | 4125(17.2) | 4875(20.3) |
| Activity physical activity, n (%) | 13,935(14.5) | 3654(15.2) | 3356(14.0) | 3535(14.7) | 3390(14.1) |
| Systolic BP, mmHg | 131±21 | 124±20 | 129±20 | 132±21 | 136±21 |
| Diastolic BP, mmHg | 83±12 | 80±11 | 83±11 | 85±1 | 87±12 |
| FBG, mmol/L | 5.47±1.67 | 4.80±0.68 | 5.09±0.76 | 5.44±1.11 | 6.54±2.66 |
| TC, mg/dL | 190.9±44.1 | 179.3±35.6 | 190.2±37.3 | 197.9±39.0 | 196.4±58.1 |
| LDL-c, mg/dL | 90.6±35.2 | 84.3±35.8 | 92.2±33.3 | 94.7±33.99 | 91.2±36.7 |
| HDL-c, mg/dL | 59.8±15.5 | 60.5±15.7 | 60.3±14.8 | 59.3±15.1 | 59.0±16.3 |
| TG, mg/dL | 112.4<br>(78.8-170.8) | 62.0<br>(51.3-72.6) | 97.4<br>(86.7-108.0) | 137.2<br>(120.4-158.4) | 245.1<br>(192.9-346.0) |
| hs-CRP, mg/dl | 0.80<br>(0.30-2.16) | 0.60<br>(0.21-1.84) | 0.72<br>(0.29-1.99) | 0.88<br>(0.33-2.16) | 1.02<br>(0.40-2.61) |
| eGFR, mL/min/1.73m<sup>2</sup> | 82.3±25.7 | 85.8±26.8 | 81.8±25.4 | 81.4±22.8 | 80.5±27.2 |
| Diabetes Mellitus, n (%) | 8408(8.8) | 223(0.9) | 542(2.3) | 1699(7.1) | 5944(24.8) |
| Hypertension, n (%) | 4,1072(42.8) | 6847(28.5) | 9651(40.2) | 11316(47.2) | 13258(55.3) |
| Anti-hypertensive drugs, n (%) | 9453(9.9) | 1440(6.0) | 1915(8.0) | 2714(11.3) | 3384(14.1) |
| Lipid-lowering drugs, n (%) | 709(0.7) | 81(0.3) | 130(0.5) | 168(0.7) | 330(1.4) |
| Diabetes drugs, n (%) | 2048(2.1) | 121(0.5) | 209(0.9) | 442(1.8) | 1273(5.3) |

Abbreviations: BMI: body mass index; BP: blood pressure; FBG: fasting blood glucose; TC: total cholesterol; LDL-c: low density lipoprotein cholesterol; HDL-c: low density lipoprotein cholesterol; TG: triglyceride; eGFR: estimated glomerular filtration rate; hs-CRP: high sensitivity C-reactive protein.

**Table 2.**
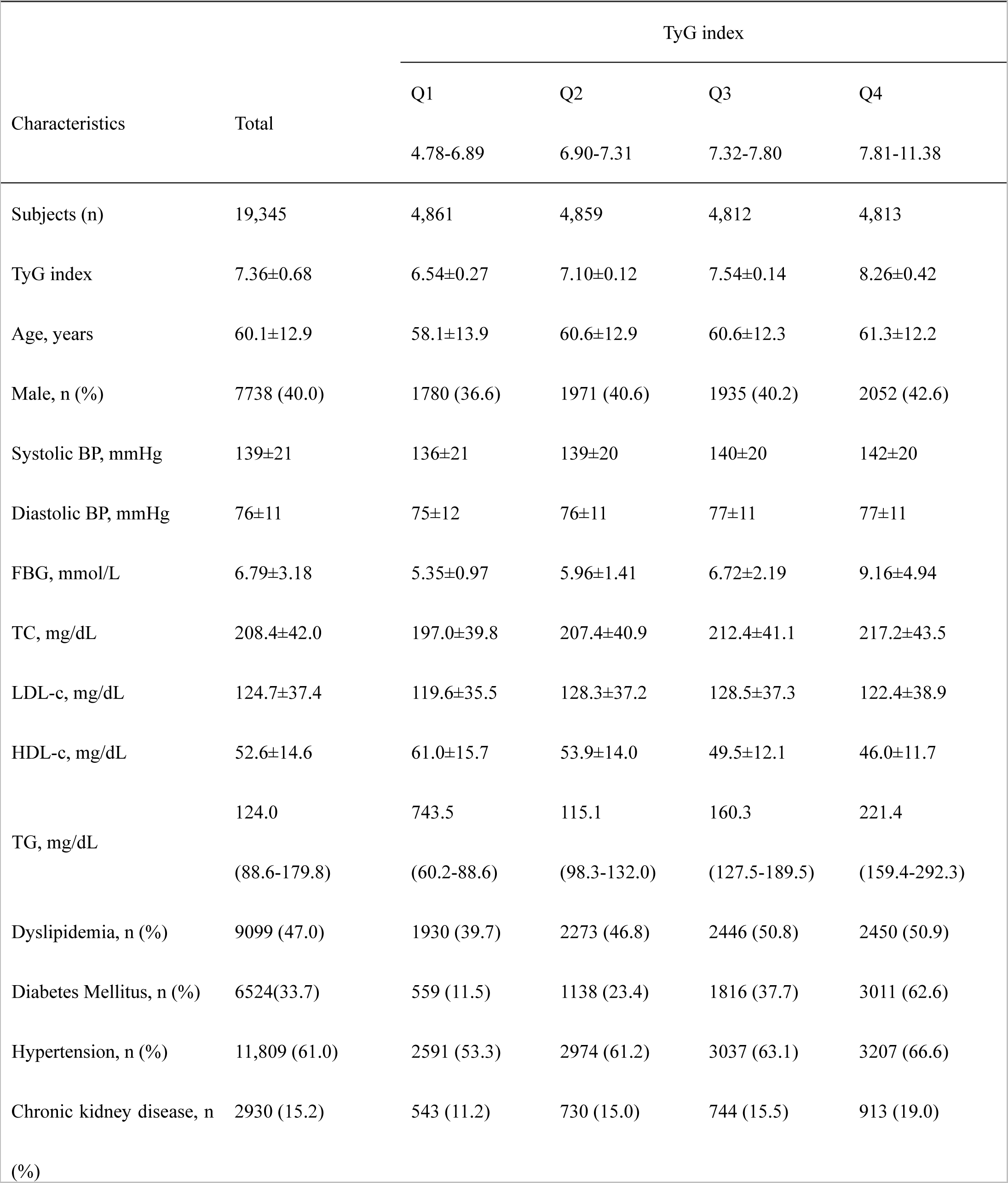
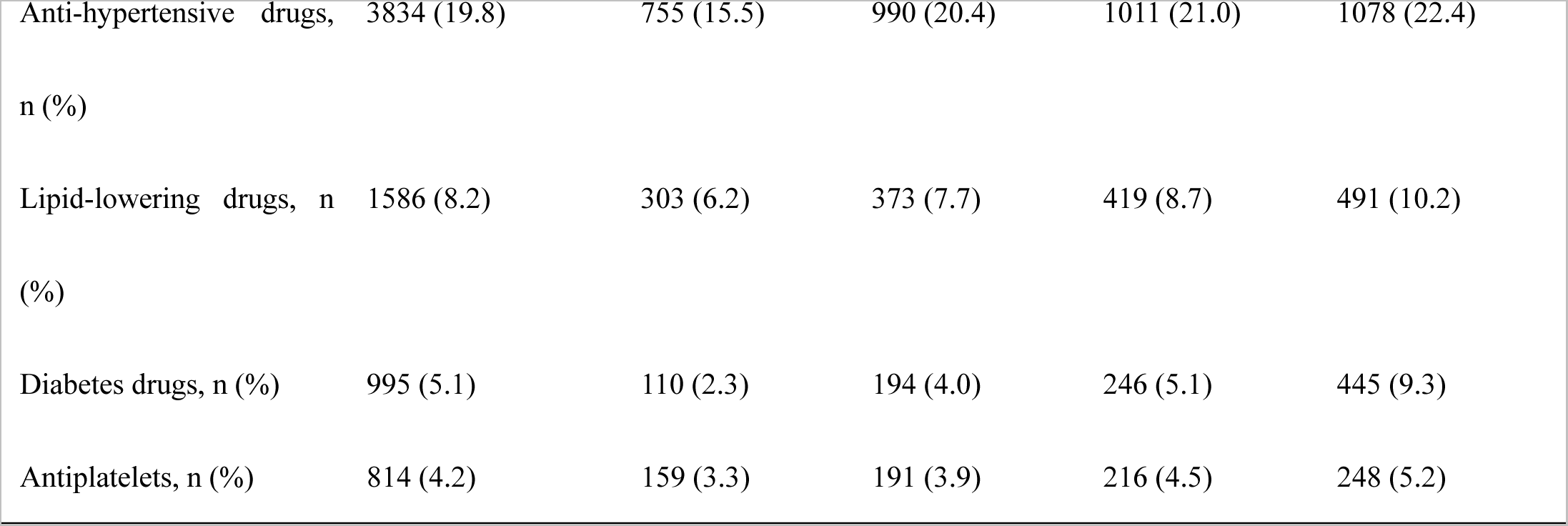
The baseline characteristics of subjects in the cohort from Hong Kong.

In the Kailuan cohort, there were 2,726 cases (2.8%) of incident HF over a mean follow-up of 12.3±2.2 years, with an overall incidence rate of 2.3 (95% CI 2.2-2.4) cases per 1000 person years. In the Hong Kong cohort, there were 1,709 cases (7.0%) of incident HF over a mean follow-up of 16.2±4.3 years, with an overall incidence rate of 5.5 (95% CI 5.3-5.8) cases per 1000 person years. Over the study duration, 10,825 subjects (11.3%) in the Kailuan cohort died (9,985 (10.1%) without developing HF), while 6,372 patients (32.9%) in the Hong Kong cohort died (4,996 (25.8%) without developing HF).

### Associations between the TyG index and the risk of incident HF

**Table 3** and **Table 4** show the associations between the TyG index, assessed both as a categorial and continuous variable, with the respective risks of incident HF in the Kailuan and Hong Kong cohorts. The cumulative incidence of incident HF for the Kailuan and the Hong Kong cohort is shown in **Figures 1A** and **1B**, respectively. After fully adjusting for potential confounders, patients in the highest quartile of the TyG index had significantly higher risks of incident HF than those in the lowest quartile in both the Kailuan (HR 1.23 (95% CI 1.09-1.39), *P* <0.001) and Hong Kong (HR 1.21 (95% CI 1.04-1.40), *P* =0.007) cohorts. Similarly, every unit increment in the TyG index was associated with a 17% and a 13% increase in the risk of HF in the Kailuan (HR 1.17 (95% CI 1.10-1.24), *P* <0.001) and Hong Kong (HR 1.13 (95% CI 1.05-1.22), *P*<0.001) cohorts, respectively. Fractional polynomial curves with full multivariable adjustment (**Figure S3**) showed a possible threshold effect in the prognostic value of the TyG index, with a lower TyG index showing no significant association with the risk of incident HF, and a higher TyG index showing a grossly linear relationship with the said risk. This was consistent with the multivariable Cox regression analysis as shown in **Tables 3 and 4** with TyG index analyzed as quartiles. Competing risk regression using the Fine and Gray sub-distribution model with death from any cause as the competing event also showed positive associations between a higher TyG index and a high risk of incident HF (**Tables 3 and 4**). Sensitivity analyses produced consistent and similar results (**Tables 3 and 4**).

**Figure 1.**
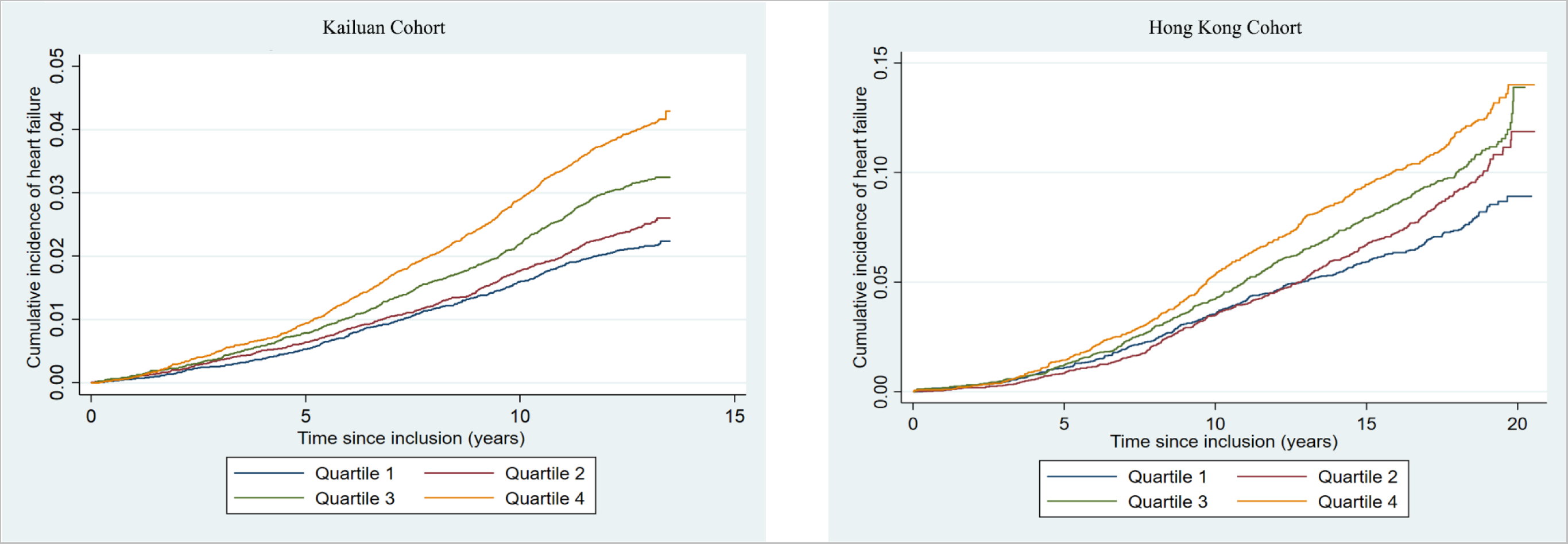
Kaplan-Meier curve of the cumulative incidence of incident heart failure for the (A) Kailuan cohort and (B) Hong Kong cohort, stratifying by quartiles of the triglyceride-glucose index.

**Table 3.** Association between baseline TyG and incident heart failure in the Kailuan cohort.

|  | Q1 | Q2 | Q3 | Q4 | *P* for trend | Per unit increment | *P* value |
| --- | --- | --- | --- | --- | --- | --- | --- |
| Number of patients | 23,997 | 24,000 | 24,001 | 23,998 |  |  |  |
| HF cases | 497 | 576 | 729 | 924 |  |  |  |
| Persons | 23997 | 24000 | 24001 | 23998 |  |  |  |
| Person-years | 297098 | 295908 | 294857 | 292552 |  |  |  |
| HF incidence <sup>1</sup> | 1.67(1.53-1.83) | 1.95(1.79-2.11) | 2.47(2.30-2.66) | 3.16(2.96-3.37) |  |  |  |
| Model 1 <sup>2</sup> | 1 | 1.16(1.03-1.31) | 1.48(1.32-1.66) | 1.90(1.70-2.11) | <0.001 | 1.44(1.37-1.51) | <0.001 |
| Model 2 <sup>3</sup> | 1 | 1.12(1.00-1.27) | 1.41(1.26-1.58) | 1.90(1.70-2.12) | <0.001 | 1.47(1.40-1.55) | <0.001 |
| Model 3 <sup>4</sup> | 1 | 1.00(0.88-1.12) | 1.12(1.00-1.26) | 1.23(1.09-1.39) | <0.001 | 1.17(1.10-1.24) | <0.001 |
| Sensitivity analysis |  |  |  |  |  |  |  |
| Sensitivity analysis 1 <sup>5</sup> | 1 | 1.00(0.88-1.13) | 1.14(1.01-1.28) | 1.23(1.10-1.41) | <0.001 | 1.18(1.10-1.25) | <0.001 |
| Sensitivity analysis 2 <sup>6</sup> | 1 | 0.97(0.84-1.11) | 1.11(0.98-1.27) | 1.17(1.02-1.34) | 0.006 | 1.13(1.06-1.22) | <0.001 |
| Competing risk regression <sup>7</sup> | 1 | 1.01(0.90-1.14) | 1.14(1.02-1.29) | 1.25(1.10-1.41) | <0.001 | 1.18(1.10-1.25) | <0.001 |

<sup>1</sup> HF incidence; The incidence rates per 1000 person years with the corresponding 95% confidence intervals are shown
<sup>2</sup> Model 1: Unadjusted. The hazard ratios with the corresponding 95% confidence intervals are shown.
<sup>3</sup> Model 2: Age-sex adjusted. The hazard ratios with the corresponding 95% confidence intervals are shown.
<sup>4</sup> Model 3: Adjusted for age, gender, education, income, physical activity, smoking status, alcohol intake, diabetes, LDL-c, HDL-c, SBP, DBP, BMI, eGFR, hs-CRP, anti-hypertensive drugs, anti-diabetes drugs, and lipid-lowering drugs. The hazard ratios with the corresponding 95% confidence intervals are shown.
<sup>5</sup> Sensitivity analysis 1: exclude follow-up time less than 2 years, remained 95,275 subjects with 2,497 HF cases. The hazard ratios with the corresponding 95% confidence intervals are shown.
<sup>6</sup> Sensitivity analysis 2: exclude subjects with medication at baseline (anti-hypertension drugs, lipid lower drugs, anti-diabetes drugs), remained 85,118 subjects with 2,118 HF cases. The hazard ratios with the corresponding 95% confidence intervals are shown.
<sup>7</sup> Competing risk regression: sub-hazard ratios with the corresponding 95% confidence intervals are shown.

**Table 4.** Association between baseline TyG index and incident heart failure in the cohort from Hong Kong.

|  | Q1 | Q2 | Q3 | Q4 | *P* for trend | Per unit increment | *P* value |
| --- | --- | --- | --- | --- | --- | --- | --- |
| Number of patients | 4,861 | 4,859 | 4,812 | 4,813 |  |  |  |
| HF cases | 342 | 404 | 454 | 509 |  |  |  |
| Persons | 4861 | 4859 | 4812 | 4813 |  |  |  |
| Person-years | 79,353 | 77,644 | 77,335 | 75,360 |  |  |  |
| HF incidence <sup>1</sup> | 4.31(3.88-4.79) | 5.20(4.72-5.74) | 5.87(5.34-6.42) | 6.75(6.19-7.37) |  |  |  |
| Model 1 <sup>2</sup> | 1 | 1.21(1.05-1.40) | 1.36(1.19-1.57) | 1.58(1.38-1.82) | <0.001 | 1.30(1.22-1.39) | <0.001 |
| Model 2 <sup>3</sup> | 1 | 1.09(0.94-1.26) | 1.23(1.07-1.42) | 1.39(1.21-1.59) | <0.001 | 1.23(1.14-1.31) | <0.001 |
| Model 3 <sup>4</sup> | 1 | 1.07(0.92-1.23) | 1.17(1.01-1.35) | 1.21(1.04-1.40) | 0.007 | 1.13(1.05-1.22) | 0.001 |
| Sensitivity analysis |  |  |  |  |  |  |  |
| Sensitivity analysis 1 <sup>5</sup> | 1 | 1.07(0.92-1.23) | 1.17(1.01-1.35) | 1.22(1.05-1.41) | 0.005 | 1.14(1.05-1.23) | 0.001 |
| Sensitivity analysis 2 <sup>6</sup> | 1 | 1.16(0.98-1.37) | 1.22(1.03-1.45) | 1.29(1.08-1.53) | 0.005 | 1.16(1.06-1.27) | 0.001 |
| Competing risk regression <sup>7</sup> | 1 | 1.06(0.91-1.22) | 1.21(1.04-1.40) | 1.24(1.07-1.44) | 0.001 | 1.15(1.06-1.24) | <0.001 |

<sup>1</sup> HF incidence; The incidence rates per 1000 person years with the corresponding 95% confidence intervals are shown
<sup>2</sup> Model 1: Unadjusted. The hazard ratios with the corresponding 95% confidence intervals are shown.
<sup>3</sup> Model 2: Age-sex adjusted. The hazard ratios with the corresponding 95% confidence intervals are shown.
<sup>4</sup> Model 3: Adjusted for age, sex, hypertension, diabetes mellitus, chronic kidney disease, dyslipidemia, antihypertensives, anti-diabetic drugs, antiplatelets, lipid-lowering drugs. The hazard ratios with the corresponding 95% confidence intervals are shown.
<sup>5</sup> Sensitivity 1: exclude follow-up time less than 2 years, remained 19,227 subjects with 1,709 HF cases. The hazard ratios with the corresponding 95% confidence intervals are shown.
<sup>6</sup> Sensitivity 2: exclude those with medications at baseline, remained 14,842 subjects with 1,223 HF cases. The hazard ratios with the corresponding 95% confidence intervals are shown.
<sup>7</sup> Competing risk regression: sub-hazard ratios with the corresponding 95% confidence intervals are shown.

Results of subgroup analyses are shown in **Figure 2A** and **Figure 2B** for the Kailuan and Hong Kong cohorts, respectively. Generally, the TyG index, analyzed as a continuous variable, was positively associated with the risk of HF across various subgroups. There was significant interaction between gender and the TyG index in the Kailuan cohort (*P* for interaction = 0.02), but not in the Hong Kong cohort (*P* for interaction = 0.11). The association between TyG index and the risk of incident HF was more prominent in female subjects than in male subjects in both cohorts [HR 1.21 (95% CI 1.02 -1.47) for female vs. 1.15 (95% CI 1.08 - 1.23) for male in the Kailuan cohort, and 1.22 (95% CI 1.10 -1.64) vs. 1.05 (95% CI 0.94 - 1.17) in the Hong Kong cohort].

**Figure 2.**
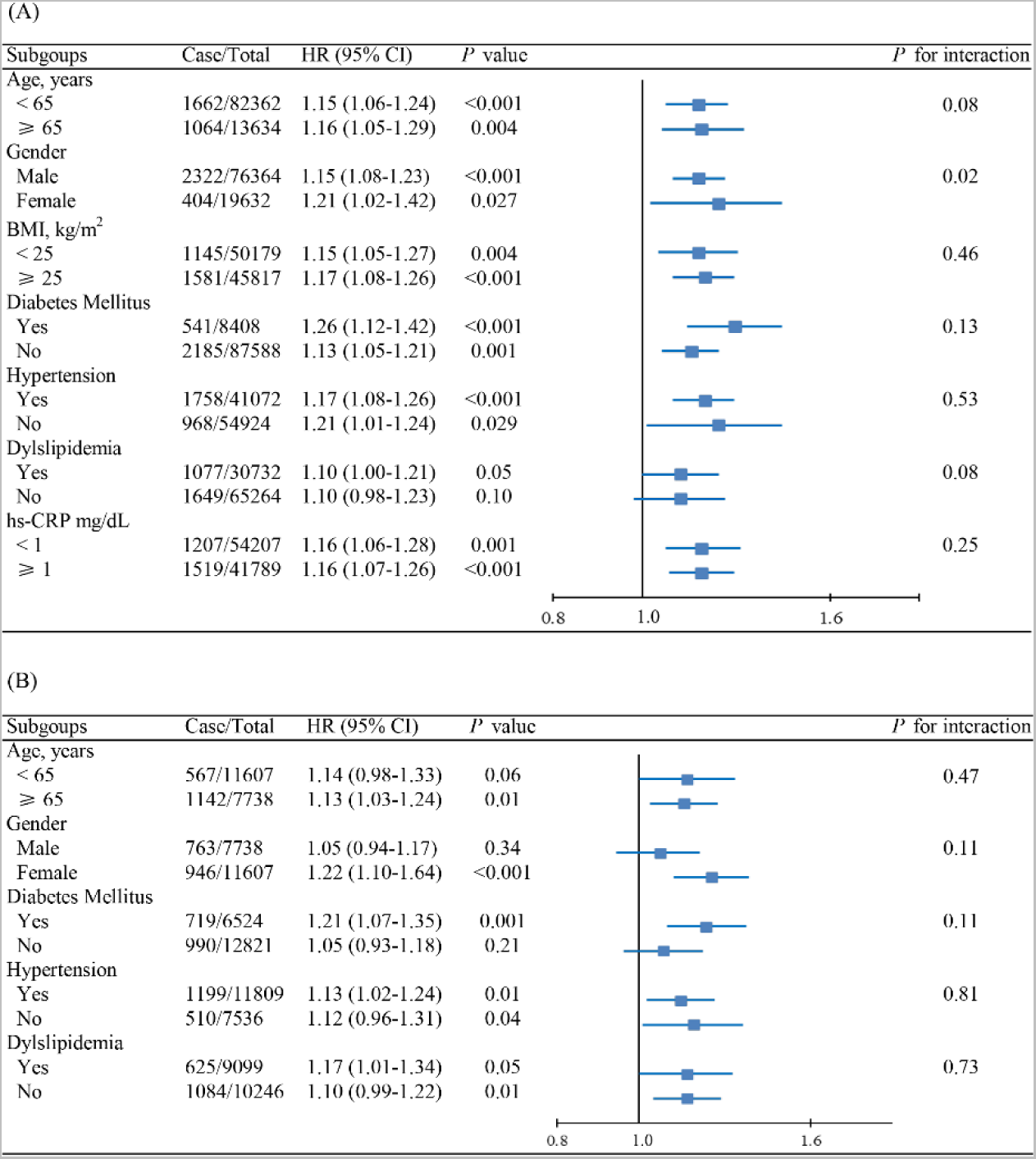
Subgroup analysis of the association between TyG index and incident HF for the (A) Kailuan cohort and (B) Hong Kong cohort. HR: hazard ratio; CI: confidence interval.

A random-effect meta-analysis combining the results from the two cohorts showed that the risk of incident HF of subjects in the highest quartile of the TyG index was 22% higher (95% CI 11% - 34%, *P*<0.0001; **Figure 3A**) than those in the lowest quartile, with every unit increment of the TyG index being associated with a 15% increase in the risk of incident HF (95% CI 10% - 21%, *P*<0.00001; **Figure 3B**). Similarly, subjects in the highest quartile of the TyG index had a 25% (95% CI 13% - 37%) increase in the sub-hazard of incident HF.

**Figure 3.**
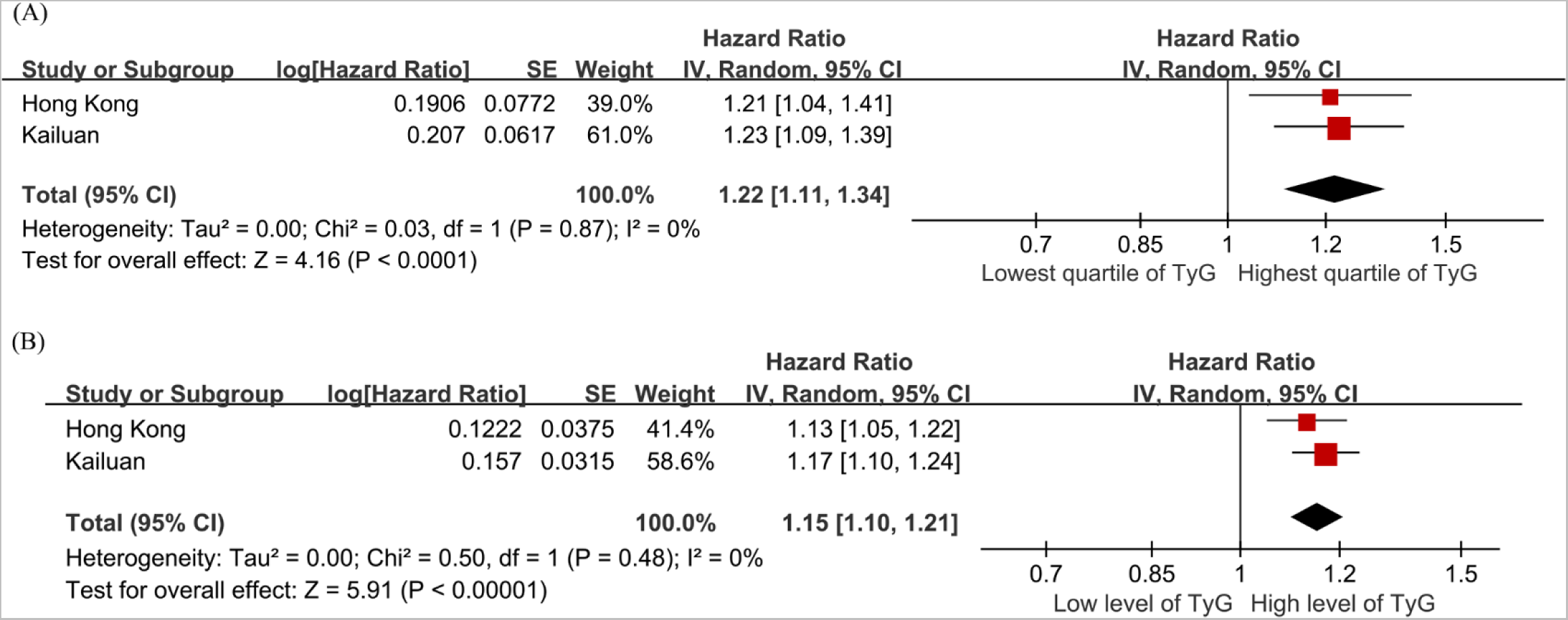
Forest plots for the meta-analysis of the association between TyG index with HF risk. (A) TyG index analyzed as a categorical variable. (B) TyG index analyzed as a continuous variable.

### Two-Sample MR analysis

The associations between genetically determined TyG index and the risk of incident HF as estimated by two-sample MR are presented in **Figure 4**. Analysis using the IVW method demonstrated that genetic predisposition to increased TyG index was significantly associated with an increased risk of incident HF (OR 1.27, 95% CI 1.15 - 1.40, *P*<0.001). The Cochran’s *Q* statistic indicated significant heterogeneity across SNPs, while no indication of directional pleiotropy was found by MR-Egger intercept (**Table S3**). The association remained consistent when using complementary methods for analysis, including weighted median, MR-Egger and MRPRESSO (**Figure 4**).

**Figure 4.**
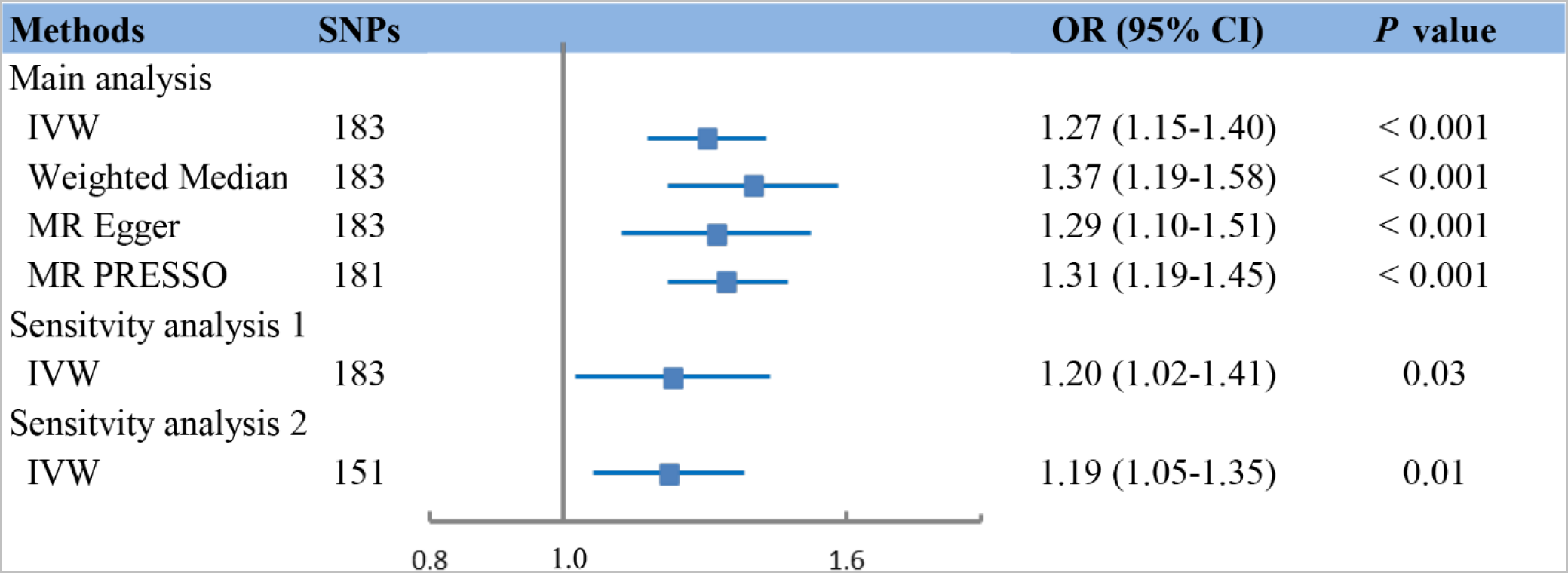
Mendelian randomization (MR) association between genetically determined TyG index and HF. Sensitivity analysis 1: multivariable MR through IVW method after adjusting for cofounders including BMI, SBP, DBP, LDL-c, and HDL-c. Sensitivity analysis 2: MR analysis through IVW after excluding any SNPs significantly associated with those confounders, including BMI, SBP, DBP, LDL-c, and HDL-c. SNPs: single-nucleotide polymorphisms; OR: odds ratio; IVW: inverse-variance weighted.

To verify the causal effect of TyG index on HF, we performed multivariable MR analysis by adjusting for HF risk factors, including BMI, blood pressure, and lipids. The association remained stable after adjusting for single risk factors (**Table S4**) and in a fully adjusted model (OR 1.20, 95% CI 1.02-1.41, *P*=0.03; **Figure 4**). Furthermore, results of the sensitivity analysis, in which 32 SNPs with potential pleiotropy were excluded, confirmed the positive association between genetically determined TyG index and HF risk (OR 1.19, 95% CI 1.05 - 1.35, *P*=0.01).

## Discussion

Utilizing observational data from two large Chinese cohorts and a two-sample MR analysis based on a public GWAS dataset, this study demonstrated that a high TyG index was an independent and causal risk factor for incident HF in the general population.

Previous studies have found independent associations between TyG index and risks of atherosclerotic cardiovascular diseases, including myocardial infarction and ischemic stroke^23, 30^. In a recent analysis of data from the Atherosclerosis Risk in Communities (ARIC) study, Huang *et al.* also reported an association between higher TyG index and higher risk of incident HF in an American population, with every standard deviation’s increase in TyG index (corresponding to a TyG index of 0.6) associated with a 15% increase in risk^31^. Our study confirmed these findings in two larger cohorts from distinct geographical regions in China. Unlike the ARIC study which was restricted to subjects between the ages of 45-64 years old, our study included adult patients across the full age range. As such, our study more closely reflects real-life practice, and our findings are thus more directly generalizable.

Importantly, utilizing MR of GWAS data, we demonstrated that the association between TyG and HF was causal by nature. Although the exact underlying mechanism for the association between TyG index and HF remains to be confirmed by further molecular studies, the well-established relationship between TyG index and insulin resistance suggests that insulin resistance may at least be an important driver of such association^10^. This was further reinforced by the results from the Kailuan cohort showing that the association between TyG index and HF was independent of chronic inflammation, as well as previous studies observing associations between insulin resistance and higher risks of incident HF independent of myocardial ischaemia^32–34^. Insulin resistance may lead to excessive circulating free fatty acids and triglycerides, which induces cardiac lipotoxicity by generating toxic lipid intermediates, and decreases cardiac efficiency by increasing fatty acid oxidation^35, 36^. Insulin resistance is also associated with disturbances of the systemic metabolic and inflammatory milieu, including increased concentrations of proinflammatory cytokines, adipokines, and catecholamines, which may trigger low-grade inflammation and chronic hypercatecholaminemia that result in detrimental effects on cardiac function^37^. Furthermore, insulin resistance is involved in the maladaptive activation of the renin-angiotensin-aldosterone system, with chronic hyperinsulinaemia inducing increased release of angiotensinogen from adipose tissue and upregulation of angiotensin II receptor expression, eventually resulting in adverse cardiac remodeling and dysfunction^38^. Nonetheless, the mechanisms between insulin resistance and HF are incompletely understood to date, and remain an important area of further research.

Another major finding of the present study is that the association between TyG index and the risk of HF was stronger in females than in males. Between-gender differences are common in cardiovascular medicine. Previous studies have shown that women with disorders of glucose metabolism have a greater risk of coronary heart disease than men^30, 39^. HF caused by obesity, diabetes, or metabolic syndrome was also found to be more common in women^40^. These suggest that between-gender differences in molecular mechanisms, particularly those in hormonal axes, may not only influence glucose and lipid metabolism, but also energy metabolism in the heart and, thereby, cardiac function.

Females are known to be less likely than males to develop insulin resistance^41^ but are at higher risk of diabetic cardiomyopathy^42^, implying that females may be more susceptible to cardiac damage induced by insulin resistance. Gender differences in nitric oxide synthase (NOS) activity and signaling, which are critical in metabolic regulation and in modulating responses to insulin resistance, are thought to be central to these observations^43^. The higher baseline levels of NOS in females predisposes to higher levels of uncoupled NOS on exposure to oxidative stress, which exacerbates the effects of insulin resistance, such as myocardial fibrosis and hypertrophy^43^. Additionally, considering that the mean age of subjects in this study implied that the female subjects were mostly postmenopausal, the postmenopausal decline in the protective effects of estrogen may contribute to gender differences in the susceptibility to insulin resistance-induced cardiac damage^40^. Notwithstanding the existing evidence as discussed above, further studies exploring the gender differences in susceptibility to insulin resistance-induced cardiac damage should provide important insights and better understanding of diabetic cardiomyopathy.

Having derived consistent findings from two geographically distinct regions in China, our results suggest that the TyG index, as a surrogate marker of insulin resistance, may be widely applicable and prognostically useful regardless of geographical region and ethnicity. As subjects with prevalent major cardiovascular diseases were excluded from the present study, the analyzed cohorts had relatively low cardiovascular risks. Our results supported the TyG index as a potentially viable and effective tool for cardiovascular risk stratification in the general population. Of note, insulin resistance in many previous studies was measured by the Homeostatic Model Assessment for Insulin Resistance (HOMA-IR) which requires measurements of fasting insulin and glucose^32, 33^. However, measuring insulin levels is expensive, and the HOMA-IR has been mostly confined to research uses with low clinical utilization. In contrast, the TyG index is simple to measure, has been validated against the euglycemic-hyperinsulinemic clamp test which is considered the gold standard for measuring insulin resistance^10^, and may outperform the HOMA-IR in identifying insulin resistance^44^. It has also been shown to be excellent at detecting insulin resistance in non-diabetic patients^45^, which is important as insulin resistance and its associated cardiovascular damage precedes overt type II diabetes mellitus^46^. The TyG index may therefore facilitate recognition of patients at elevated risk of incident HF, for which efficacious measures for primary prevention exist^4^.

### Strengths and limitations

The strengths of our study included the large sample size, long follow-up time, and having demonstrated reproducible results across two independent observational cohorts and MR analysis. Our findings were further strengthened by multiple subgroup and sensitivity analyses yielding largely consistent results. To the best of the authors’ knowledge, this was one of the first studies demonstrating causality between higher TyG index and higher risk of incident HF. Nonetheless, some limitations must be noted. First, we were unable to compare the predictive power of different methods for assessing insulin resistance in our observational study, since fasting insulin levels were unavailable for most subjects. Second, inherent to all observational studies, there may be residual or unmeasured confounders that we were not able to address. Nonetheless, we have included multiple important risk factors for incident HF in the multivariable regression models, and the numerous sensitivity analyses yielded consistent results which reinforced the validity of our findings. Third, the MR analysis was restricted to patients of European descent to reduce bias from population stratification, which may limit extrapolation of our MR results to other populations. Nevertheless, given that associations between TyG index and the risk of incident HF observed in a recent report in an American cohort (the ARIC study) were comparable to our findings as observed in Chinese cohorts, the causality established by our MR analysis is likely true in Chinese patients as well. Fourth, no information was available about the subtype of incident HF. Given the different metabolic mechanisms contributing to the pathogenesis of different types of HF^47^, further research in this regard is warranted. Fifth, diagnoses of the Hong Kong cohort were identified using ICD-9 codes and could not be individually adjudicated due to the retrospective, deidentified nature of the database, as well as the large sample size. Regardless, all diagnostic codes were entered by treating clinicians, who were completely independent of the authors. CDARS has also been shown to have good coding accuracy, specifically for cardiovascular outcomes^20^.

## Conclusion

As observed from two large, geographically distinct Chinese cohorts, a higher TyG index was independently associated with higher risk of incident HF. MR analysis demonstrated that the association was likely causal in nature. Further studies are warranted to confirm our findings and fully elucidate the underlying biological mechanisms.

## Data Availability

All data produced in the present study are available upon reasonable request to the authors

## Acknowledgements

The authors appreciate all the subjects involved in this study, their families, and the members of the survey team from the Kailuan community.

## Author contributions

Xintao Li and Jeffrey Shi Kai Chan designed this study, conducted the main analysis, visualized the results, and wrote the first draft of the manuscript. Bo Guan, Shi Peng, Xiaoyu Wu, Yan Hiu Athena Lee, Jeremy Man Ho Hui, Danish Iltaf Satti, Shek Long Tsang, Shouling Wu, Jiandong Zhou, Songwen Chen, Gary Tse, and Shaowen Liu reviewed and edited the article. All authors contributed to the article significantly and approved the submitted version.

